# Liquid Plasma vs Thawed Plasma: Tracking Coagulation Factor Activity Changes During Storage

**DOI:** 10.1101/2025.08.13.25333231

**Authors:** Nalan Yurtsever, Catherine Gereg, Nichelle Perera, Parveen Bahel, Melissa Alicea, Henry M. Rinder, Edward L. Snyder, Christopher A. Tormey, Edward S. Lee

## Abstract

**Background and Objectives:** Liquid plasma (LQP) stands out as an alternative to thawed plasma (TP) for emergent transfusions due to longer shelf life. We aim to measure fibrinogen, Protein C, Protein S, FV, FVII, and FVIII activity in LQP, quantify how these factors levels change during storage, and characterize how they compare in LQP to TP.

**Materials and Methods:** Coagulation factor activities were measured on Days 15, 26, and 27 for LQP (n=26) and Day 5 for TP (n=31). Bayesian statistics was used to compare coagulation factor activity and quantify changes in activity during storage.

**Results:** Fibrinogen and Protein C activity in Day 26 LQP (LQP26) was comparable to Day 5 TP (TP5) with posterior mean activity of 257 mg/dL vs 246 mg/dL and 100.4% vs 108.7%, respectively. FV, FVII, and FVIII had lower activity in LQP26 vs TP5 with posterior mean activities of 42.6% vs 72.0%, 55.0% vs 59.7%, and 48.8% vs 59.2%, respectively. Protein S in LQP26 was low with posterior mean activity of 28.0%, which was less than half that of TP5 at 66.4%. From Day 15 to Day 26, FVII in LQP decreased at a rate of -3.49% per day whereas fibrinogen, Protein C, Protein S, FV, and FVIII activity in LQP remained relatively stable.

**Conclusion:** Compared to TP5, LQP26 has comparable activities of fibrinogen, Protein C, FVII, lower activities of FV and Protein S, and slightly lower activity of Factor VIII. LQP is a viable alternative for use in emergency transfusions and massive transfusion protocols.

**Highlights:**

– Liquid plasma has comparable activities to thawed plasma for fibrinogen, Protein C, and Factor VII, which is advantageous for emergency use due to its extended shelf life.
– Liquid plasma has adequate fibrinogen and Protein C levels on the last day of expiration.
– Liquid plasma has ∼50% FV, FVII and FVIII activity levels on the last day of expiration, making it a sufficient replacement option to TP for active bleeding.

## Introduction

Administration of plasma has an important role in acute trauma care as early transfusion of red blood cells, plasma, and platelets in a 1:1:1 ratio is recommended during resuscitation [1]. Despite increased usage of low titer whole blood in the trauma setting (PMID: 38483205), component therapy remains as the standard of care in most institutions. In the United States, plasma components can be stored frozen for long periods of time (e.g., 1 year at -18 °C or colder) but require thawing before use. Once thawed, plasma has a shelf life of 24 hours stored at 1–6 °C, and after this initial 24-hour post-thaw period, plasma can be stored up to four more days as “thawed plasma” (TP). These storage requirements cause blood banks to face challenges in maintaining an active plasma inventory for rapid release. Consequently, thawed plasma units are routinely wasted to keep an inventory of plasma products immediately available for emergent indications. However, this approach can still cause delays in issuing plasma if all thawed plasma units are employed and frozen plasma components need to be thawed.

Liquid plasma (LQP) is an alternative to TP and frozen plasma products for rapid availability of plasma for resuscitation. This never-frozen product is stored at 2–6 °C and has a shelf life of 26 days, allowing for increased flexibility in inventory management and fast utilization in massive transfusion protocols [2, 3]. As such, one study investigating wastage after switching from a TP-only inventory to a mixed TP and LQP inventory observed a drop in yearly wastage rates from 13.5% to 10.3%.(PMID: 33105307) Another study showed that implementation of LQP resulted in a significant improvement in 24-hour plasma:RBC transfusion ratios from 1:1.72 to 1:1.23 and observed reductions in hospital length of study, 24-hour mortality, and acute kidney injury [4]. Multiple other retrospective studies showed no difference between TP and LQP for length of stay and survival [5-8].

Yet, there has been limited evaluation of the hemostatic profile of LQP. One study performed thromboelastogram measurements of TP and LQP and found significantly higher maximum amplitude, shear elastic modulus strength, and total thrombus generated in LQP, which was partly attributed to increased residual platelets in LQP [9]. Another study suggested that the leukocyte and platelet counts in all plasma products are similar but that cells are disrupted due to freezing and thawing, resulting in unreliable measurements in frozen plasma products such as fresh frozen plasma (FFP) and TP. (PMID: 31029763) In addition, during storage, LQP had significant loss of labile coagulation factors such as factor V, factor VIII, and protein S, but other factors maintained stable levels. LQP is also shown to be as effective as TP in blocking endothelial permeability [10].

In this study, we aim to characterize how select coagulation factor levels change in liquid plasma units during storage. We performed serial measurements in LQP units at mid-expiration (Day 15), on the day of expiration (Day 26), and the day after expiration (Day 27). Day 27 measurements were included to see if any changes in coagulation factor activity occur one day after expiration. Additionally, we compare coagulation factor levels in LQP to TP on the final day of storage (Day 5) to compare the hemostatic profile of these plasma products.

## Materials and Methods

### Measurement of coagulation factors in liquid plasma and thawed plasma

We obtained 26 units of liquid plasma and 31 units of FFP from the American Red Cross (Farmington, Connecticut, United States). Liquid plasma units were all blood group A, and thawed plasma units were group A (n=20), O (n=9), or AB (n=2). All plasma units were derived from blood collected in citrate-phosphate-dextrose. All LQP units were irradiated upon arrival at our blood bank by an X-ray irradiator (Gilardoni S.p.A. Radgil-2, Italy), which was done to prevent transfusion associated graft versus host disease (TA-GVHD) that could be precipitated by viable leukocyte that are typically inactivated by the freezing process in frozen plasma products. To obtain units of thawed plasma at Day 5 (TP5), FFP was thawed in a water bath at 37°C according to standard AABB operating procedures and stored at 1–6°C for 5 days. Aliquots of LQP were obtained from the same units on Day 15, Day 26 (day of expiration), and Day 27 (1 day after expiration). Day 15 samples were obtained by using segments included with the unit, and Day 26 and 27 samples were obtained by using sterile connections to obtain aliquots of approximately 15 mL from the units (Terumo TSCD-11 Sterile Tubing Welder, Terumo BCT, Highlands Rach, Colorado, U.S.). Additionally, to determine if coagulation factor activity in segments and in bags correlate, we collected paired segments and aliquots from bags using sterile connections from 2 units of liquid plasma and 10 units of thawed plasma. These samples were then frozen and stored at −70°C until time of testing of coagulation factors. Aliquots or segments of TP5 were similarly obtained at Day 5 of storage and were frozen and stored at −70°C until time of testing. On the day of testing, aliquots were thawed at 37°C in a water bath. Samples that looked unclear or turbid were centrifuged at 10,000 rpm × 10 minutes (Hettich RotoFix32 centrifuge, Beverly, MA) to remove any debris.

We measured the activity level of Factor V (FV), Factor VII (FVII), and Factor VIII (FVIII) to account for common, extrinsic, and intrinsic pathways along with fibrinogen to account for the final step of plasma hemostasis. We also measured Protein C (PC) and Protein S (PS) activities. FV, FVII, FVIII, PC, and PS activities were tested using a clinical-grade analyzer (ACL TOP 750, Bedford, MA) based on prothrombin time correction of factor deficient plasma and expressed as percent activity. Fibrinogen was tested using a quantitative clot-based analyzer (Siemens CS-5100, Malvern, PA). The normal range for human plasma factor levels at our facility is established by reference range studies of normal individuals. This study was reviewed by the Yale University Institutional Review Board and was determined to not constitute human subjects research.

### Statistical Analysis

We used Bayesian statistics for our statistical analysis due to several advantages, which includes flexibility in modeling longitudinal data, incorporation of prior information, and validity for any sample size. We estimated the average coagulation factor activities in LQP at Day 15 (LQP15), LQP at Day 26 (LQP26), LQP on Day 27 (LQP27), and TP5 using the Bayesian analog of the one-sample t-test, and we estimated the difference in factor activities between LQP26 and TP5 using a Bayesian version of the two-sample t-test [11]. Full mathematical details for these statistical models are described in Supplementary Information (SI) Section S2 and Section S3 respectively.

To characterize the change in coagulation factor activity over storage time, we implemented multilevel piecewise linear regression models adapted from a previously described approach to model how factor activity levels change with storage time [12]. These models estimate the rate of change per storage day from Day 15 to Day 25 and the rate of change per day from Day 26 to Day 27. This structure provides a comparison of the magnitude of coagulation factor level changes between these two time periods of storage. The multilevel regression structure captures the longitudinal information in the repeated time point measurements for every plasma unit, allowing for inferences in factor level changes for each individual plasma unit and for the whole population of plasma units. Complete description of the statistical approach is described in SI, Section S4.

We compared the coagulation factor levels between TP5 units obtained from donors of different ABO types. Data from type A and AB TP5 units were combined and compared to type O TP5 units using the Bayesian two-sample t-test (SI Section S5). To test the correlation in coagulation factor activity between samples from segments and units, we performed linear regression between paired segment and unit factor activities (SI Section S6).

We used weakly informative or informative priors, which that were motivated by previously described factor levels in other plasma products [13]. Details about priors are included in SI, Sections S2–S4. The Hamiltonian Monte Carlo engine Stan (v2.36) was used to estimate posterior distributions of each model with R (v4.4.3) using CmdStanR (v0.8.1) and rethinking packages (v2.42) [14-16]. Statistical models were either coded directly in the Stan language or with the rethinking package using the ulam function. Each model was run with 4 Hamilton Monte Carlo chains of 10,000 samples per chain in parallel with half as warmup until convergence achieved as suggested by high effective sample sizes, R-hat estimates of 1.00, and visual inspections of traceplots. Bayesian statistical inference does not return point estimates for parameters but instead provides posterior distributions for possible parameter values, and posterior distributions are summarized with the posterior mean values and 95% credible intervals (95% CI). Data and code implementing the models are available on github.com/edwardslee/liquidplasma.

## Results

We analyzed the activities of fibrinogen, PC, PS, FV, FVII, and FVIII in 26 LQP units on Days 15 (except for FV), 26, and 27 of storage and in 31 TP5 units (Figure 1A). We compared the activity levels of these factors in paired aliquots from bags and segments and found that measurements correlated between these two sample sources except for FV (SI Section S6). The posterior mean estimates for the average coagulation activity with 95% credible intervals are shown in Table 1, and observed descriptive statistics are shown in Tables S1–S2. The posterior mean factor activity in LQP15 for fibrinogen was 241 mg/dL (95% CI: 228, 254 mg/dL), for PC was 110.6% (95% CI: 102.8%, 118.7%), for PS was 35.3% (95% CI: 27.1%, 43.6%), for FVII was 66.1% (95% CI: 55.0%, 80.0%), and for FVIII was 56.0% (95% CI: 51.8%, 60.2%). Since all LQP15 samples were measured in segments, we could not perform this analysis for FV activity for LQP15. Posterior summaries for factor activities for LQP26, LQP27, and TP5 are shown in Table 1.

**Table 1.**
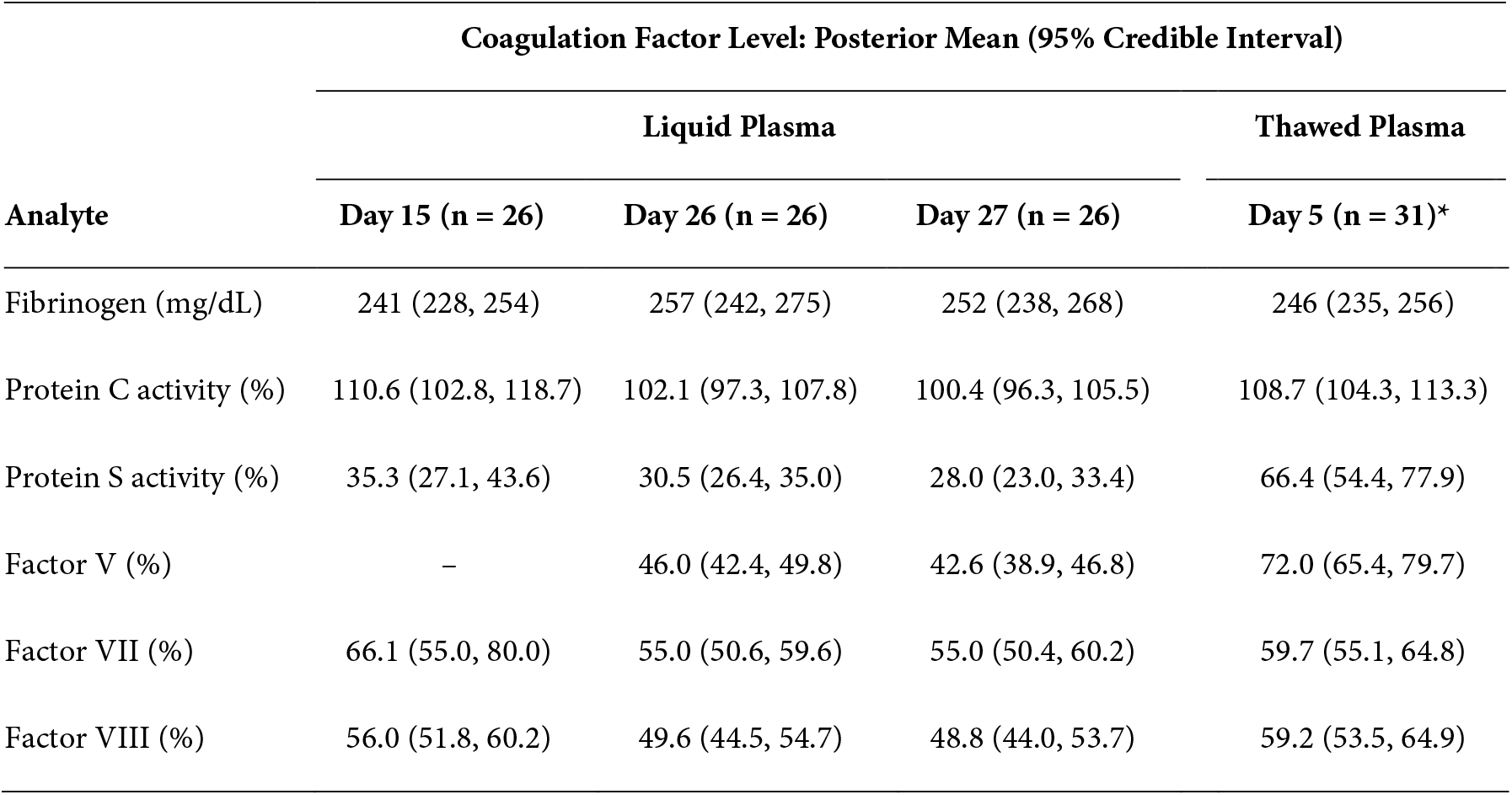
Average coagulation factor activity in liquid plasma and thawed plasma. Estimation of mean coagulation factor activity of fibrinogen, Protein C, Protein S, Factor V, Factor VII, and Factor VIII levels in liquid plasma at Day 15 of storage, liquid plasma at Day 26 of storage (day of expiration), liquid plasma at Day 27 of storage (1 day of expiration), and thawed plasma at Day 5 of storage (day of expiration). The table shows the posterior mean estimates with 95% credible intervals in parentheses. *n = 8 for Factor V.

| Analyte | Coagulation Factor Level: Posterior Mean (95% Credible Interval) |  |  |  |
| --- | --- | --- | --- | --- |
|  | Liquid Plasma |  |  | Thawed Plasma |
|  | Day 15 (n = 26) | Day 26 (n = 26) | Day 27 (n = 26) | Day 5 (n = 31)* |
| Fibrinogen (mg/dL) | 241 (228, 254) | 257 (242, 275) | 252 (238, 268) | 246 (235, 256) |
| Protein C activity (%) | 110.6 (102.8, 118.7) | 102.1 (97.3, 107.8) | 100.4 (96.3, 105.5) | 108.7 (104.3, 113.3) |
| Protein S activity (%) | 35.3 (27.1, 43.6) | 30.5 (26.4, 35.0) | 28.0 (23.0, 33.4) | 66.4 (54.4, 77.9) |
| Factor V (%) | – | 46.0 (42.4, 49.8) | 42.6 (38.9, 46.8) | 72.0 (65.4, 79.7) |
| Factor VII (%) | 66.1 (55.0, 80.0) | 55.0 (50.6, 59.6) | 55.0 (50.4, 60.2) | 59.7 (55.1, 64.8) |
| Factor VIII (%) | 56.0 (51.8, 60.2) | 49.6 (44.5, 54.7) | 48.8 (44.0, 53.7) | 59.2 (53.5, 64.9) |

**Figure 1.**
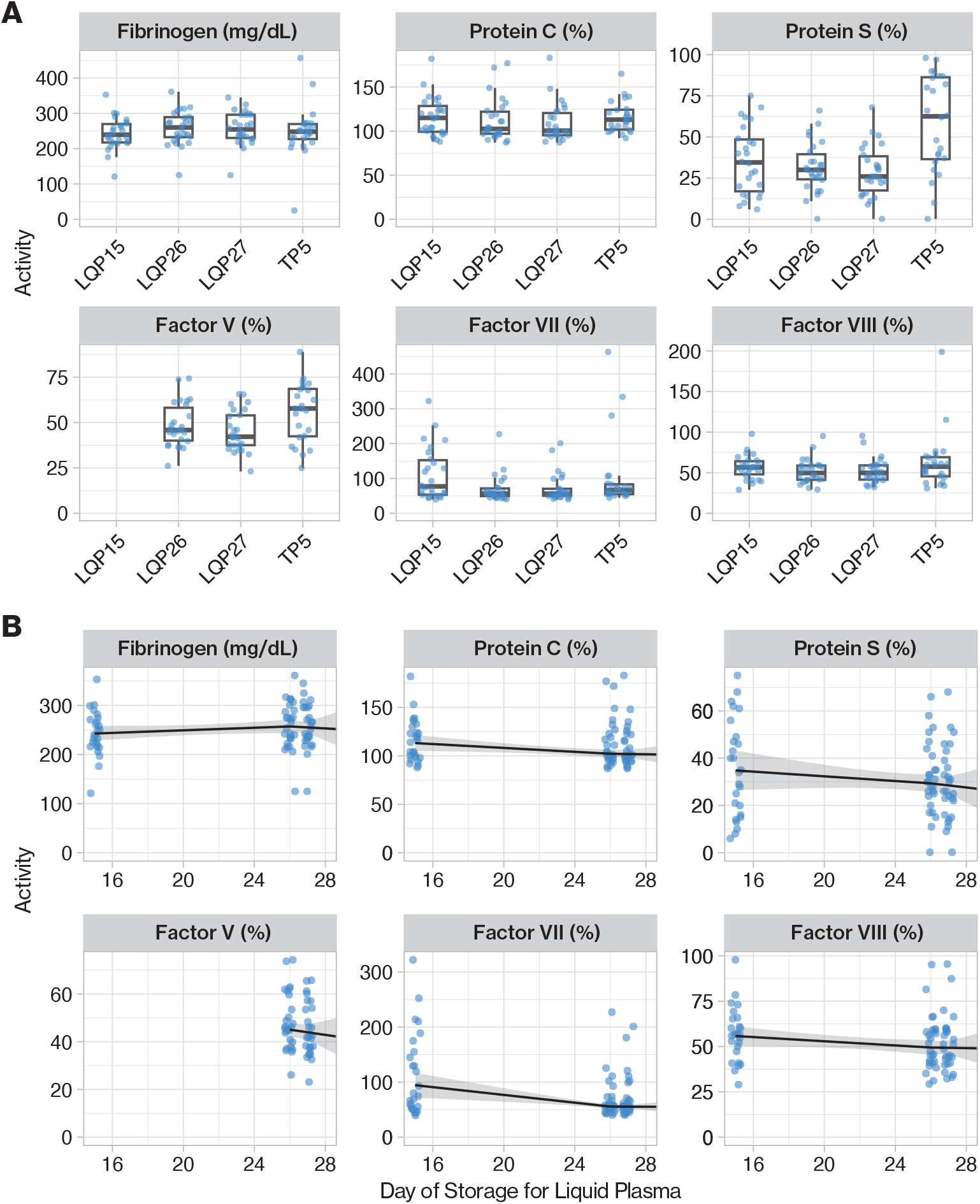
Coagulation factor activity in liquid plasma and thawed plasma. (A) Coagulation factor activity levels of fibrinogen, Protein C, Protein S, Factor V, Factor VII, and Factor VIII were measured in liquid plasma units on Day 15 of storage (LQP15), on Day 26 of storage (LQP26), and on Day 27 of storage (LQP27), which is one day after expiration. The same coagulation factors were measured in thawed plasma units on Day 5 of storage. The box-and-whisker plots depict the minimum, first quartile (25^th^ percentile), median, third quartile (75^th^ percentile), and maximum values. (B) Multilevel piecewise linear regression was performed for all coagulation factors in liquid plasma units to characterize how coagulation factor activity changes with storage time. Plots depict posterior mean fits with 95% credible interval envelopes with individual factor activity measurements as jittered points.

We compared LQP26 and TP5 to characterize the differences in coagulation factors at expiration for these two plasma products (Table 2). The differences in fibrinogen, PC, and FVII have wide 95% credible intervals that contain 0 and thus support that LQP26 and TP5 have similar levels of these coagulation factors. In contrast, the posterior mean difference between LQP26 and TP5 for PS was -33.6% (95% CI: -47.7%, -18.8%) and for FV was -27.1% (95% CI: -38.9%, -16.7%), providing strong evidence that PS and FV is substantially lower in LQP26 than in TP5. The posterior mean difference for FVIII was -9.4% with a 95% CI of -17.8% to -1.0%, suggesting that FVIII is lower in LQP26 and TP5 but with a smaller magnitude than PS or FV.

**Table 2.** Difference in coagulation factor activity between LQP26 and TP5 and changes in factor activity during storage for liquid plasma. Posterior mean estimates are shown with 95% credible intervals in parentheses.

| Analyte | Difference in activity<br>between LQP26 and TP5 | Change in factor activity per day in liquid plasma |  |
| --- | --- | --- | --- |
|  |  | Day 15 to Day 26 | Day 26 to Day 27 |
| Fibrinogen (mg/dL) | 13.6 (-8.2, 35.5) | 1.3 (-0.57, 3.18) | -2.0 (-16.8, 12.9) |
| Protein C (%) | -5.5 (-13.6, 3.24) | -1.0 (-1.8, -0.18) | -0.26 (-3.8, 3.2) |
| Protein S (%) | -35.4 (-48.4, -21.9) | -0.5 (-1.3, 0.32) | -0.88 (-4.2, 2.6) |
| Factor V (%) | -27.1 (-38.9, -16.7) | – | – |
| Factor VII (%) | -4.9 (-13.1, 3.4) | -3.5 (-5.5, -1.4) | -0.17 (-3.6, 3.2) |
| Factor VIII (%) | -9.4 (-17.8, -1.0) | -0.6 (-1.2, 0.04) | -0.18 (-3.6, 3.3) |

We quantified the changes in coagulation factor activity during storage in liquid plasma units using multilevel piecewise linear regression models that estimate the average rate of change per day from mid-expiration (Day 15) to expiration (Day 26) and estimate the average rate of change from expiration to one day after expiration (Day 27). The posterior mean rates of change and 95% credible intervals of these parameters are shown in Table 2.

FVII demonstrated a considerable decrease during the Day 15 to Day 26 storage period with a posterior mean rate of change of -3.5% per day (95% CI: - 5.5%, -1.4%). The other coagulation factors had rates of changes from Day 15 to Day 26 with small effect sizes and credible intervals close to 0, indicating strong statistical evidence for small magnitudes of change in factor activity during storage compared to the large decrease observed for FVII. Fibrinogen showed minimal increase during storage with a posterior mean rate of change of 1.31 mg/dL per day (95% CI: -0.57, 3.18) with a credible interval containing 0 and strongly indicating a negligible effect size. Similarly, PC, PS, and FVIII had small decreases during storage posterior mean rates of change of -1.00% per day (95% CI: -1.83, -0.18), -0.50% per day (95% CI: -1.34, 0.32), and -0.58% (95% CI: -1.19, 0.04), respectively, with credible intervals that are close to or include 0.

From Day 26 to Day 27, fibrinogen, PC, PS, FVII, and FVIII had small posterior mean rates of change with wide 95% credible intervals that include 0. These posterior distributions strongly support that these coagulation factors do not significantly change one day after expiration and indicate that changes in coagulation factors occur substantially before expiration during storage.

Since all the LQP units were group A and the TP5 samples were a mix of A, AB, and O, we addressed a potential limitation that coagulation factor activity comparisons can be confounded by ABO group differences while noting that a majority of the TP5 samples were group A. We found no strong evidence of differences in any coagulation factors between A/AB and O TP5 units, with all 95% CIs including 0 (SI Section 5, Table S3), and our results indicate that our LQP and TP5 samples can be directly compared.

## Discussion

Liquid plasma offers an extended refrigerated shelf life, which enables immediate use, compared to thawed plasma while retaining most essential coagulation factors needed for acute and emergent resuscitation needs. Our study evaluated fibrinogen, FV, FVII, FVIII, PS, and PC in a considerable number of LQP units during days of storage that would be typically found in inventory for blood banks and compared the activity of these coagulation factors to thawed plasma. We found that LQP15 has high fibrinogen and protein C activities with mean levels within 1 standard deviation of previously described levels in FFP and PF24 at thaw but contains levels of protein S, FVII, and FVIII more than 1 standard deviation lower than those described in FFP at thaw [11,21]. We found that LQP15 similarly has lower protein S than PF24 but FVII and FVIII within or close to 1 standard deviation within levels described in PF24 at thaw. These results are generally consistent with previously published findings for liquid plasma as Matijevic et al. observed similarly high activity of fibrinogen and protein C and lower levels of protein S, FV, and FVIII, and Meledeo et. al found similar lower levels of FV and FVIII [9, 17].

The findings of our comparison between LQP and TP strongly support that LQP is a viable alternative to thawed plasma for acute plasma resuscitation needs. The mean activities of fibrinogen, FVII, and PC in LQP26 remained comparable to TP5 and are at levels similar to previously reported values for thawed FFP at 120 hours post thaw [18-20]. In contrast, we found that PS and FV levels are considerably lower in LQP26 than in TP5 and that FVIII activity in LQP26 is lower than in TP5 with a smaller magnitude. The mean activities for PS and FVIII in LQP26 are more than 1 standard deviation lower than reported levels found in FFP at 120 hours post thaw, and the mean activity for FVII in LQP26 is similar to those found in FFP [11,21]. These results demonstrate that LQP cannot substitute FFP in non-emergent settings. However, fibrinogen loss is one of the major concerns in acute trauma settings, and since fibrinogen activities are similar in LQP and TP5, LQP can be used as a readily available plasma product for acute resuscitation indications when there is not enough time to thaw frozen plasma products [17]. In addition, FVII has comparable activity in LQP and TP5 with an average activity of 55.0% in LQP27, which is sufficient for immediate coagulation [21].

Although there are no quality controls requirements for coagulation factor levels in liquid plasma products in the United States or in Europe to our knowledge, the Council of Europe and European Directorate for the Quality of Medicines & HealthCare (EDQM) have defined specifications for fresh frozen plasma products.[22] EDQM has requirements for FFP to have an average FVIII activity of at least 70 IU/100 mL after thaw, for pathogen reduced FFP to have an average activity of at least 50 IU/100 mL after thaw, and for at least 90% of units to have FVIII activity of at least 50 IU/100 mL for both. Our results show that neither LQP on Day 15, 26, or 27 nor TP5 meet the 70 IU/100mL average requirement for FFP (Table S1 and S2) and that 73% of LQP15 units (19/26) and 71% of TP units (22/31) have FVIII activity greater than or equal to 50 IU/100mL, thus not meeting EDQM requirements. In contrast, both LQP and TP meet the requirement for pathogen reduced FFP to have a minimum average FVIII activity of 50 IU/100 mL. These results are not surprising due to the lability of FVIII, but our findings suggest similarities in FVIII to pathogen reduced plasma with average content meeting requirements and ∼70% of units containing specified minimum FVIII activity. These comparisons further support the usage of LQP as an alternative for TP in acute indications for plasma transfusion.

The piecewise linear regression analysis demonstrated that FVII decreases during storage whereas fibrinogen, FVIII, PC, and PS either do not change or change to a small degree from Day 15 to Day 27. These results are consistent with FVII being a labile coagulation factor in plasma products but then suggest that the other four coagulation factors can be considered stable in liquid plasma during this period of storage. Our conclusions about FVII partially agree with one study that observed an initial decline in FVII in the first 10–15 days of storage and then a subsequent increase in FVII but disagrees with another study that saw an increase in FVII during storage [9, 23]. However, the effect size and significance of the findings from these respective studies are unclear as Matijevic et al. had descriptive statistics without statistical modeling and Gosselin et al. used second-order polynomial regression without a clear casual interpretation. In addition, methodological differences could explain some of these discrepancies as our study and Gosselin et al. analyzed samples that were frozen and thawed while Matijevic et al. tested fresh samples.

One major limitation in our study was our approach for sampling segments of units. To obtain longitudinal factor activity data at multiple time points, we decided to sample segments from liquid plasma units on Day 15. Our correlation analysis between factor activities in segments and aliquots from the unit bag showed that all our measured coagulation factors correlated well except for FV, which showed lower levels in segments. As a result, we could not include FV activities for LQP15 in our analysis, and only TP5 samples that were aliquoted were analyzed. This observed discordance between segment and bag FV activities could be due to the known lability of FV and warrants further investigation.

In our hospital system, LQP units are initially delivered to the main hospital, where they are irradiated and then distributed to the smaller satellite and community hospitals in the system. Irradiation is performed to prevent the possibility of TA-GVHD due to the viable leukocytes present in LQP and is unlikely to affect coagulation factors. By the time LQP reaches inventory in these other hospitals, they are typically 10–15 days old, and LQP units that are issued can be anywhere from 15 to 26 days old. Although we found that PS and FV are lower in LQP26 compared to TP5, we also found that the most important hemostatic coagulation factors, particularly fibrinogen and FVII, are similar in LQP26 and TP5. These findings help to justify using primarily LQP in our hospital system as an alternative to TP for acute resuscitation needs while allowing for more flexibility for managing plasma inventory, resulting in a decrease in our system-wide plasma wastage from 17% to 13%.

We aim that the results of the comparison between LQP26 and TP5 can provide guidance in resources about plasma components, such as the Circulation of Information [13].

The stability of the coagulation factors from Day 26 to Day 27 seen in our study warrants more investigation if the shelf life of liquid plasma can be extended beyond 26 days. One preclinical study observed that the activity levels of FV and FVIII significantly decreased from Day 0 to Day 40 of storage but saw stable activity of fibrinogen throughout this period. The observed decrease in this study likely occurred in the first two weeks of storage and thus does not conflict with our results that FVIII and other labile coagulation factors are stable after Day 15. Additionally, clinical data show no significant difference in patient outcomes between those receiving older versus newer LQP units, supporting this perspective [7]. Further studies measuring additional coagulation factors at later time points will be needed to confirm the longevity of LQP, but our study suggests that liquid plasma could be extended at least one day past expiration.

Our study provides more evidence that liquid plasma is a viable alternative to thawed plasma as an effective strategy to quickly provide plasma products for initial emergency resuscitation while allowing sufficient time for thawing frozen plasma. LQP provides both volume expansion and coagulation support while significantly reducing blood product wastage due to its longer shelf life compared to thawed plasma.

## Data Availability

The data supporting the findings of this study are publicly available and can be downloaded from github.com/edwardslee/liquidplasma

## Acknowledgements

N.Y., C.G., N.P., E.S.L., C.A.T., H.M.R. designed the study. N.Y., C.G., N.P., E.S.L. collected samples and maintained the study database. P.B. and M.A. analyzed the samples in the lab. C.A.T., E.L.S., and H.M.R. supervised the study. N.Y. and E.S.L. analyzed the data. N.Y., C.G. and E.S.L. wrote the manuscript. All authors reviewed and approved the manuscript.

We thank all the blood bank staff at Bridgeport Hospital, Milford Campus, and Yale New Haven Hospital and the hematology staff at Yale New Haven Hospital for their help in this project.

