## Supplementary Information for "Liquid Plasma vs Thawed Plasma: Tracking Coagulation Factor Activity Changes During Storage"

### Supplemental Information for "Liquid Plasma vs Thawed Plasma: Tracking Coagulation Factor Activity Changes During Storage"

Nalan Yurtsever, Catherine Gereg, Nichelle Perera, Parveen Bahel, Melissa Alicea, Henry M. Rinder, Edward L. Snyder, Christopher A. Tormey, Edward S. Lee

#### S1 Descriptive Statistics

Descriptive statistics for factor activity data are shown for liquid plasma on Days 15 (LQP15), 26 (LQP26), and 27 (LQP27) in Table S1 and for thawed plasma on Day 5 (TP5) in Table S2. Since LQP15 units were sampled with segments, we could not use FV measurements due to lack of consistent correlation with bag activities (see Section S6

| | Day 15 | | | Day 26 | | | Day 27 | | |
|---|---|---|---|---|---|---|---|---|---|
| Analyte | Mean | SD | Range | Mean | SD | Range | Mean | SD | Range |
| Fibrinogen (mg/dL) | 243.0 | 45.6 | 121–353 | 258.7 | 48.0 | 125–361 | 257.1 | 46.3 | 125–345 |
| Protein C (%) | 116.2 | 22.1 | 88–182 | 112.5 | 24.1 | 87–177 | 109.5 | 21.7 | 87–183 |
| Protein S (%) | 35.6 | 20.4 | 6–75 | 31.7 | 15.1 | 0.2–66 | 29.0 | 15.6 | 0.2–68 |
| Factor V (%) | – | – | – | 48.6 | 12.0 | 26.1–74.2 | 45.0 | 11.2 | 23.1–65.7 |
| Factor VII (%) | 112.5 | 75.6 | 39.6–322.2 | 69.1 | 39.0 | 40–227.1 | 72.7 | 41.0 | 40.1–201.1 |
| Factor VIII (%) | 56.6 | 14.8 | 29–97.8 | 51.8 | 15.0 | 29.3–95.2 | 52.1 | 15.5 | 32.2–95.5 |

Table S1: Descriptive statistics for liquid plasma coagulation factor activity measurements

| | Day 5 | | |
|---|---|---|---|
| Analyte | Mean | SD | Range |
| Fibrinogen (mg/dL) | 259 | 85.8 | 25–531 |
| Protein C (%) | 112 | 18.2 | 75.0–165 |
| Protein S (%) | 61.3 | 30.2 | 0.2–106.0 |
| Factor V (%) | 78.8 | 15.6 | 63.8–88.9 |
| Factor VII (%) | 92.3 | 93.4 | 37.1–463.1 |
| Factor VIII (%) | 61.0 | 30.9 | 30.9–198.8 |

Table S2: Descriptive statistics for thawed plasma coagulation factor activity measurements

#### S2 Student-t distribution model

We used a Student-t distribution model as a Bayesian analog to the frequentist one-sample t test to estimate the average coagulation factor activity in LQP15, LQP26, LQP27, and in TP5. Here, we choose to use the Student-t distribution instead of the Normal distribution since the thicker tails of the Student-t distribution reduce the influence of extreme values, which can be seen in factor activity measurements. As such, this approach can be thought of as a Bayesian analog of the one-sample t test.

For a given coagulation factor and day of storage, let $f_i$ be the activity for liquid plasma unit $i$. The activity measurements have a Student-t distribution with mean $\mu$ and standard deviation $\sigma$

$$f_i \sim \text{Student}(2, \mu, \sigma)$$

where we fix the shape parameters $\nu$ to be 2. The shape parameter is empirically fixed to be 2 since letting $\nu$ to be free generally results in worse or equal out-of-sample predictive accuracy (discussed below in Section S7). The priors were motivated by values listed for FFP and TP in the Circular of Information for the Use of Human Blood and Blood Components, and the same priors were used for LQP15, LQP26, and LQP27 [1].

For fibrinogen, the following priors were used:

$$\mu \sim \text{Normal}(280, 50)$$
$$\sigma \sim \text{Exponential}(1).$$

For Protein C, the following priors were used:

$$\mu \sim \text{Normal}(90, 20)$$
$$\sigma \sim \text{Exponential}(1).$$

For Protein S, the following priors were used:

$$\mu \sim \text{Normal}(80, 20)$$
$$\sigma \sim \text{Exponential}(1).$$

For Factor V, the following priors were used:

$$\mu \sim \text{Normal}(60, 20)$$
$$\sigma \sim \text{Exponential}(1).$$

For Factor VII, the following priors were used:

$$\mu \sim \text{Normal}(70, 20)$$
$$\sigma \sim \text{Exponential}(1).$$

For Factor VIII, the following priors were used:

$$\mu \sim \text{Normal}(50, 20)$$
$$\sigma \sim \text{Exponential}(1).$$

#### S3 Bayesian two-sample t test

We adapted a Bayesian version of the two-sample t test by Kruschke to compare the mean coagulation factor activity between LQP26 and TP5 [3]. The model uses the Student-t distribution to describe the distribution of factor activity measurements with different means $\mu_{\text{Product}[i]}$ and standard deviations $\sigma_{\text{Product}[i]}$ for Product$[i] \in \{$LQP26, TP5$\}$, and as in Section S2, we fix $\nu$ to be 2. Unlike Kruschke's approach, priors for $\mu_i$ and $\sigma_i$ are not estimated from the group mean and standard deviations. Instead, priors for $\mu_i$ are the same as those used in the models described in Section S2, and weakly informative exponential priors were used for $\sigma_i$. The full model specifications are

$$f_i \sim \text{Student}\big(2, \mu_{\text{Product}[i]}, \sigma_{\text{Product}[i]}\big)$$
$$\mu_j \sim \text{Priors as described above}$$
$$\sigma_j \sim \text{Exponential}(0.2).$$

The posterior difference in mean factor activity between LQP26 and TP5 was calculated with the difference $\mu_{\text{LQP26}} - \mu_{\text{TP5}}$.

#### S4 Bayesian Piecewise Linear Regression

We aimed to estimate the change in coagulation factor activity during storage, and we particularly wanted to make inferences on the effect sizes of the rate of change from Day 15 to Day 26, the rate of change from Day 26 and Day 27, and the difference between the two change during these two periods. These would allow us to conclude how much coagulation factor activity changes during storage and if the changes differ from Day 15 (mid-expiration) to Day 26 (expiration) and from expiration to Day 27 (one day after expiration).

In order to do so, we employed Bayesian multilevel piecewise linear regression models, which are adapted from the approach described by Brilleman et al. with several changes [2]. In our model, we use a "robust" regression approach using a Student-t likelihood instead of the usual Normal likelihood since the thicker tails of the Student-t distribution reduces the influence of extreme values, which are often observed in factor activity measurements. We also fix the "change point" of the piecewise linear regression model to be Day 26, which is the day of expiration of liquid plasma. Finally, the multilevel structure models the individual random effects for each LQP unit and use partial pooling to estimate the population level rate of change in factor activity.

The data are modeled with a Student-t likelihood with $\nu$ fixed at 2 as before with a piecewise linear mixed effects model with the form

$$
\begin{aligned}
f_i &\sim \text{Student}(2, \mu_i, \sigma) \\
\mu_i &= \alpha + \beta_{\text{Unit[i]}}(t_i - 26)I(26 - t_i) + \gamma_{\text{Unit[i]}}(t_i - 26)I(t_i - 26) \\
\sigma_i &\sim \text{Exponential}(1) \\
\alpha &\sim \text{Priors as described below} \\
\beta_j &\sim \text{Normal}(\bar{\beta}, \sigma_\beta) \\
\bar{\beta} &\sim \text{Priors as described below} \\
\sigma_\beta &\sim \text{Exponential}(1) \\
\gamma_j &\sim \text{Normal}(\bar{\gamma}, \sigma_\gamma) \\
\bar{\gamma} &\sim \text{Priors as described below} \\
\sigma_\gamma &\sim \text{Exponential}(1)
\end{aligned}
$$

where $I(.)$ is the indicator function such that

$$
I(26 - t_i) = \begin{cases} 1 & \text{if } t_i < 26 \\ 0 & \text{if } t_i \geq 26 \end{cases} \qquad I(t_i - 26) = \begin{cases} 0 & \text{if } t_i < 26 \\ 1 & \text{if } t_i \geq 26 \end{cases}
$$

Here, we fixed the change point for the piecewise linear regression to be 26, which is the day of expiration for liquid plasma. The priors for $\alpha$ are the same priors used for $\mu$ in Section S2, and the priors for $\bar{\beta}$ and $\bar{\gamma}$ were chosen to be weakly informative.

For fibrinogen, the following priors were used:

$$
\begin{aligned}
\bar{\beta}, \bar{\gamma} &\sim \text{Normal}(0, 10) \\
\alpha &\sim \text{Normal}(280, 50).
\end{aligned}
$$

For Protein C, the following priors were used:

$$
\begin{aligned}
\bar{\beta}, \bar{\gamma} &\sim \text{Normal}(0, 2) \\
\alpha &\sim \text{Normal}(90, 20).
\end{aligned}
$$

For Protein S, the following priors were used:

$$
\begin{aligned}
\bar{\beta}, \bar{\gamma} &\sim \text{Normal}(0, 2) \\
\alpha &\sim \text{Normal}(80, 20).
\end{aligned}
$$

For Factor V, the following priors were used:

$$\bar{\beta}, \bar{\gamma} \sim \text{Normal}(0, 2)$$
$$\alpha \sim \text{Normal}(60, 20).$$

For Factor VII, the following priors were used:

$$\bar{\beta}, \bar{\gamma} \sim \text{Normal}(0, 2)$$
$$\alpha \sim \text{Normal}(70, 20).$$

For Factor VIII, the following priors were used:

$$\bar{\beta}, \bar{\gamma} \sim \text{Normal}(0, 2)$$
$$\alpha \sim \text{Normal}(70, 20).$$

Note that the code implementing these models have Day 15 set to $t = 0$, and the priors for $\bar{\beta}$ and $\bar{\gamma}$ use a non-centered parameterization to increase sampling efficiency and to avoid divergent transitions with the Stan Hamiltonian Monte Carlo engine.

#### S5 Comparison of coagulation factors of different ABO types in thawed plasma samples

Although all liquid plasma samples were derived from group A donors, TP5 samples were from a mix of type A (n = 20), AB (n = 2), and O (n = 9) donors. Since some studies have shown that type O individuals have lower Factor VIII and von Willebrand factor levels, we wanted to address any potential confounding that could be caused by this mix of ABO types by comparing coagulation factor activities group O TP5 units with group A and AB TP5 units using the Bayesian two-sample t test approach described in Section S3 (described in more detail below).

We found no strong evidence suggestive of differences in any of the coagulation factors between group O and non-group O TP5 units, with all 95% credible intervals including 0 (Table S3). To further confirm these results, we also performed the frequentist one-way ANOVA procedure to compare coagulation factor activity across group O, group A, and group AB TP5 units and found no statistically significant differences in all coagulation factors using the conventional $p$ value cutoff of 0.05 ($p > 0.05$).

Letting $f_i$ be the factor activity for TP5 unit $i$, we would like to estimate mean activity $\mu_{\text{ABO}[i]}$ with standard deviation $\sigma_{\text{ABO}[i]}$, where ABO[$i$] $\in \{A/AB, O\}$. The full model specifications are below.

| | Posterior Mean Difference | 95% CI |
|---|---|---|
| Fibrinogen (mg/dL) | -12.9 | -30.7, 6.10 |
| Protein C (%) | 1.56 | -10.1, 13.4 |
| Protein S (%) | -11.6 | -23.4, 1.34 |
| Factor V (%) | -7.32 | -16.4, 2.33 |
| Factor VII (%) | -10.4 | -21.8, 1.71 |
| Factor VIII (%) | 7.13 | -7.35, 20.6 |

Table S3: Comparison of coagulation factor activity between group A/AB thawed plasma units and O thawed plasma units. All 95% credible intervals include 0, suggesting no large difference between these two groups of plasma units.

For fibrinogen:

$$
\begin{aligned}
f_i &\sim \text{Student}\big(2, \mu_{\text{ABO}[i]}, \sigma_{\text{ABO}[i]}\big) \\
\mu_j &\sim \text{Normal}(280, 10) \\
\sigma_j &\sim \text{Exponential}(0.2)
\end{aligned}
$$

For Factor V:

$$
\begin{aligned}
f_i &\sim \text{Student}\big(2, \mu_{\text{ABO}[i]}, \sigma_{\text{ABO}[i]}\big) \\
\mu_j &\sim \text{Normal}(60, 5) \\
\sigma_j &\sim \text{Exponential}(0.2)
\end{aligned}
$$

For Factor VII:

$$
\begin{aligned}
f_i &\sim \text{Student}\big(2, \mu_{\text{ABO}[i]}, \sigma_{\text{ABO}[i]}\big) \\
\mu_j &\sim \text{Normal}(80, 5) \\
\sigma_j &\sim \text{Exponential}(0.2)
\end{aligned}
$$

For Factor VIII:

$$
\begin{aligned}
f_i &\sim \text{Student}\big(2, \mu_{\text{ABO}[i]}, \sigma_{\text{ABO}[i]}\big) \\
\mu_j &\sim \text{Normal}(50, 20) \\
\sigma_j &\sim \text{Exponential}(0.2)
\end{aligned}
$$

For Protein C:

$$
\begin{aligned}
f_i &\sim \text{Student}\big(2, \mu_{\text{ABO}[i]}, \sigma_{\text{ABO}[i]}\big) \\
\mu_j &\sim \text{Normal}(90, 10) \\
\sigma_j &\sim \text{Exponential}(0.2)
\end{aligned}
$$

For Protein S:

$$\begin{aligned} f_i &\sim \text{Student}\left(2, \mu_{\text{ABO}[i]}, \sigma_{\text{ABO}[i]}\right) \\ \mu_j &\sim \text{Normal}(70, 5) \\ \sigma_j &\sim \text{Exponential}(0.2) \end{aligned}$$

#### S6 Determining the correlation between bag and segment coagulation factor activity

We wanted to determine if fibrinogen, Factor V (FV), Factor VII (FVII), Factor VIII (FVIII), protein S (PS), and protein C (PC) activity measurements correlated between segments and aliquots from the unit bag. We measured the activities of these coagulation factors in paired segment and aliquot samples from 2 units of liquid plasma and 10 units of thawed plasma. We then performed linear regression to see how well segment values predicted unit bag values with the following general form

$$\begin{aligned} f_i^{\text{bag}} &\sim \text{Normal}(\mu_i, \sigma) \\ \mu_i &= \beta f_i^{\text{segment}} + \alpha \\ \beta, \alpha &\sim \text{Priors below} \\ \sigma &\sim \text{Exponential}(0.2). \end{aligned}$$

Here, for each unit $i$, $\beta$ is the slope that correlates segment factor activity $f_i^{\text{segment}}$ with bag factor activity $f_i^{\text{bag}}$.

Mean posterior estimates and 95% credible intervals (CIs) are shown in Table S4, and mean posterior fits with 95% CI envelopes are shown in Figure S1. Our results strongly support that segment and bag activities correlate well for fibrinogen, FVII, FVIII, PS, and PC since slopes are close to 1 and intercepts are close or near 0. Although the regression results show some potential minor bias towards lower values in segments for PS, the posterior distributions for the slope and intercept parameters are close enough to 1 and 0 respectively to not significantly impact accuracy. However, factor V does not correlate well as segment factor V activity is lower than bag activity with an average shift of ~27%. As a result, segments do not reliably predict FV activity in unit bags but are accurate for the other coagulation factors.

The following priors were used. The priors for fibrinogen are

$$\begin{aligned} \beta &\sim \text{Normal}(1, 0.2) \\ \alpha &\sim \text{Normal}(0, 20) \end{aligned}$$

| | Slope | Intercept |
|---|---|---|
| Fibrinogen | 0.94 (0.83, 1.05) | 24.62 (-1.82, 51.15) |
| Protein C | 1.01 (0.96, 1.06) | 1.84 (-3.34, 6.78) |
| Protein S | 1.13 (1.03, 1.23) | 3.25 (-1.71, 8.08) |
| Factor V | 0.70 (0.38, 1.03) | 26.61 (7.36, 44.88) |
| Factor VII | 1.05 (0.99, 1.1) | -2.84 (-6.82, 1.21) |
| Factor VIII | 0.96 (0.80, 1.12) | 4.51 (-6.87, 15.63) |

Table S4: Posterior distributions of linear regression between factor activity levels in bags and segments. These results indicate that all coagulation factors correlate well between segments and bags except for factor V, which shows higher levels in bags compared to segments. Posterior means are shown with 95% credible intervals in parentheses.

The priors for Factor V are

$$\beta \sim \text{Normal}(1, 0.25)$$
$$\alpha \sim \text{Normal}(0, 20)$$

The priors for Factor VII and Factor VIII

$$\beta \sim \text{Normal}(1, 0.1)$$
$$\alpha \sim \text{Normal}(0, 20)$$

The priors for Protein S are

$$\beta \sim \text{Normal}(1, 0.08)$$
$$\alpha \sim \text{Normal}(0, 3)$$

The priors for Protein C are

$$\beta \sim \text{Normal}(1, 0.1)$$
$$\alpha \sim \text{Normal}(0, 5)$$

Figure S1: Coagulation factor activity was measured in paired segments and aliquots from liquid plasma and thawed plasma units. Mean posterior regression line with 95% CI envelopes are shown.

#### S7 Comparing using fixed $\nu$ to setting $\nu$ free in Student-t likelihoods

All of the models described in Sections S2–S4 use a fixed value of 2 for the shape parameter $\nu$. We evaluated the out-of-sample accuracy setting $\nu$ fixed to 2 compared to setting $\nu$ to be free using wAIC and generally found that using fixed $\nu$ was favored. For example, if we fit the following two models to the fibrinogen data in LQP26,

$$\begin{aligned} f_i &\sim \text{Student}(2, \mu, \sigma) \\ \mu &\sim \text{Normal}(280, 50) \\ \sigma &\sim \text{Exponential}(1) \end{aligned}$$

and

$$\begin{aligned} f_i &\sim \text{Student}(\nu, \mu, \sigma) \\ \mu &\sim \text{Normal}(280, 50) \\ \sigma &\sim \text{Exponential}(1) \\ \nu &\sim \text{Exponential}(1/10) \end{aligned}$$

we get the following results in Table S5, which shows that the fixed $\nu$ model is clearly favored with a model weight of 1 using wAIC. Thus, to simplify the models, we use $\nu = 2$ for all the models in this study. The full analysis for all time points and coagulation factors are in the script `student-t-analysis.R` in the Github repository.

| Model | wAIC | Weight |
|---|---|---|
| Fixed $\nu$ | 294.5 | 1 |
| Free $\nu$ | 312.7 | 0 |

Table S5: Comparison of Student-t models with a fixed $\nu$ (set at 2) and free $\nu$ for fibrinogen in LQP26.
